# Gender-affirming hormone therapy and impacts on quality of life: a narrative review

**DOI:** 10.1101/2025.03.11.25323442

**Authors:** Logan Powell, Anahi Puebla, Rebecca J Lepping

**Affiliations:** Department of Neurology, University of Kansas Medical Center, 3901 Rainbow Blvd, Kansas City, KS USA; Hoglund Biomedical Imaging Center, University of Kansas Medical Center, 3901 Rainbow Blvd, Kansas City, KS, USA; University of Kansas School of Medicine, University of Kansas Medical Center, 3901 Rainbow Blvd, Kansas City, KS, USA

**Keywords:** transgender, gender-affirming care, mental health, quality of life, healthcare disparities

## Abstract

**Background:** Transgender and gender-nonconforming (TGNC) people often face significant disparities in health education and access to quality medical management. This narrative literature review examines the relationship between TGNC patients seeking hormone replacement therapy and resulting improved health outcomes.

**Methods:** Our search identified papers through the databases PubMed, PsycINFO, CINAHL, Embase, and Web of Science including search terms relating to gender-affirming hormone therapy (GAHT), transgender identities, and patient healthcare experiences and outcomes. Further inclusion criteria required papers published after 1979 with a majority of participants located in the United States. Data extraction and quality assessment of the selected papers were completed using the JBI Manual for Evidence Synthesis, a quality assessment tool created based on the Mixed Methods Appraisal Tool, and Covidence software. Common themes were narratively reviewed.

**Results:** The search yielded 19,482 results across five databases and 51 papers were included in data extraction and quality assessment. Most papers were published between 2020-2024 and enrolled young adults in cross-sectional studies. Recurrent themes observed from data synthesis include improved mental health and quality of life outcomes associated with GAHT use. Distance to clinics, cost of care, insurance coverage, and governmental policies were commonly identified barriers to obtaining gender-affirming care.

**Conclusions:** The identified gaps in information reflect the importance of additional research in TGNC health-related disparities including diverse participant populations and rigorous longitudinal methods. With these changes, we expect improved quality of care, patient satisfaction, and health outcomes for these individuals.

**HIGHLIGHTS:**

- Gender-affirming hormone therapy is associated with reduced levels of depression, anxiety, and suicidal ideation in transgender and gender-nonconforming individuals
- Significant barriers to obtaining GAHT include high costs, lack of insurance coverage, limited access to knowledgeable healthcare providers, geographical distance to clinics, and discriminatory policies
- The number of studies on gender-affirming hormone therapy has increased significantly in recent years, reflecting growing recognition of the importance of transgender healthcare
- Most existing studies on GAHT and its effects are cross-sectional, limiting the ability to assess long-term outcomes
- Establishing standardized assessments for mental health outcomes, quality of life, and long-term effects of gender-affirming hormone therapy would enhance the reliability and comparability of future research

---

Transgender and gender-nonconforming (TGNC) people, like other marginalized groups in healthcare, often face significant disparities in health outcomes and access to quality medical management surrounding the unique needs of these patients. While many TGNC people seek care for gender-affirming hormone therapy (GAHT) and gender-affirming surgeries, previous research identifies that this population’s health is also disproportionately affected by risk factors such as mental illness, substance abuse, and HIV, requiring comprehensive healthcare beyond gender-affirming interventions alone.^1^ Inconsistent health outcomes for TGNC people have frequently been attributed to factors such as income inequality, limited access to health insurance, lack of knowledgeable healthcare providers, and experiences of direct discrimination by medical professionals.^2^ The 2022 U.S. Transgender Survey of 92,329 participants reported that approximately one-quarter of TGNC participants have avoided medical treatment due to concerns about financial costs or fear of discrimination.^3^ With a larger percent of today’s population identifying as transgender and seeking gender-affirming care, the need for addressing these disparities becomes even more important.^4^

Many studies have identified a need for increasing the number of knowledgeable healthcare providers prepared to care for TGNC patients.^1,5,6^ The shortage of education on the care of this population ultimately discourages both providers from offering care and patients from seeking it.^1,2,7^ Previous research suggests that health outcomes for this population could improve significantly with increased support for the dissemination of standards of care guidelines and the development of “both trans-focused and trans-friendly primary care models.”^7^ Building on these findings, this study seeks to explore the relationship between access to GAHT, a common first step in medical transition, and improved healthcare engagement and outcomes.

Our research aims to highlight the importance of expanding transgender healthcare access and education, with the goal of preparing future healthcare providers to address the needs of TGNC patients with appropriate expertise and cultural humility. In this paper we present a narrative review of published literature that examines the relationship between TGNC patients seeking hormone replacement therapy and resulting improved health outcomes as well as the barriers they face in receiving this care. As this is a relatively new area of research, our review aims to evaluate the current body of literature, identify gaps in knowledge to guide future research, and reduce healthcare disparities and promote equal health outcomes for TGNC people.

## METHODS

### Search Strategy

A structured search strategy identified papers through the databases PubMed (NCBI), PsycINFO (EBSCO), CINAHL Complete (EBSCO), Embase (Elsevier), and Web of Science Core Collections - SCI-Expanded 1900-present, SSCI 1956-present, AHCI 1975-present, ESCI 2020-present (Clarivate). The search strategy consisted of three categories of search terms: gender-affirming hormone therapy (GAHT), transgender or gender-nonconforming identities, and patient healthcare experiences and outcomes. These three categories were connected by the “AND” Boolean operator while terms in each category were linked by the “OR” operator. Relevant papers and published sexual minority search criteria were reviewed to identify search terms.^8^ While some of the terms used are considered outdated or offensive to some individuals, they have been included to broaden the results of database searches and account for the terminology used in older papers. A summarized example of the search terms is shown in Table 1, while the full search strategies can be found in Supplemental Materials A. Controlled vocabulary such as MeSH and Emtree terms were included when available. No other limits were applied to the search.

**Table 1.** Abbreviated list of terms used for title and abstract search in literature review to analyze GAHT impact on quality of life.

| **Gender-affirming hormone therapy (connected by OR)** |  | **Transgender and gender-nonconforming identity terms (connected by OR)** |  | **Patient healthcare experiences and outcomes (connected by OR)** |
| --- | --- | --- | --- | --- |
| Gender affirming hormon* |  | LGBT* |  | Patient compliance |
| Hormone replacement therapy |  | transgender* |  | Patient adherence |
| HRT |  | Transvestite |  | Quality of life |
| Puberty blockers |  | Transsexual |  | Psychological well-being |
| Puberty inhibitors | **AND** | crosssex* | **AND** | Cultural competenc* |
| Gender-affirming care |  | FTM |  | Treatment outcome |
| Cross sex hormon* |  | MTF |  | Mental health |
| Feminizing hormon* |  | Nonbinary |  | Behavioral health |
| Masculinizing hormon* |  | trans* people |  | Self-esteem |
| Testosterone block* |  | trans* man |  |  |
| Endocrinology |  | trans* woman |  |  |
|  |  | Gender dysphoria* |  |  |
|  |  | Gender non-conforming |  |  |
|  |  | Gender identity |  |  |
|  |  | Gender reassignment |  |  |

*Terms were searched for in titles and abstracts across all databases used. Asterisks indicate wildcard truncations, searching for any variation of the listed word. Full search strategies can be found in Supplemental Materials A.*

Titles and abstracts were searched, and results were uploaded into the Covidence software (Veritas Health Innovation Ltd, Melbourne). The search was completed on May 22^nd^, 2024, and a total of 19,840 citations were uploaded to Covidence. While this search strategy resulted in a large and broad collection of papers that required additional screening of irrelevant titles, we felt it was essential to capture all papers that included the diverse vocabulary used for identifying and affirming TGNC people.

### Paper Selection

After importing the search results from each database into Covidence, duplicate papers were identified and removed through both an automated and manual process. The remaining paper title and abstracts were screened for relevance based on key inclusion criteria (Table 2). Papers were limited to studies conducted primarily in the U.S. and published in English as the experience of TGNC people is often unique to the political and cultural climate of their country of residence. In addition, papers were eliminated if published before 1979, when the first standards of care for transgender patients were published by WPATH.^9^ All remaining peer-reviewed, research papers in reference to the social and emotional impacts of hormone replacement therapy on TGNC patients and the barriers to affirming care they experience were further reviewed. This process of title and abstract screening was completed by a single reviewer (R1), and excluded papers that did not meet these criteria.

**Table 2.** Inclusion and exclusion criteria used for paper selection in literature review to analyze GAHT impact on quality of life.

| **Inclusion Criteria** | **Exclusion Criteria** |
| --- | --- |
| **Pediatric, adolescent, and adult populations** | Newborn and infant populations |
| **Population identifies as transgender or gender nonconforming** | Does not reference the transgender population/participants do not identify as transgender |
| **Human participants** | Non-human participants |
| **Majority of participants sought or used gender-affirming hormone therapy (testosterone, estrogen, spironolactone, hormone blockers, puberty inhibitors/blockers)** | Majority of participants have NOT desired or used gender-affirming hormone therapy |
| **Results discuss the psychosocial impacts of taking or barriers to GAHT** | Exclusively discusses physical/metabolic impacts of GAHT |
| **Published DURING or AFTER 1980** | Published BEFORE 1980 |
| **Majority of study participants located WITHIN the United States** | Majority of study participants located OUTSIDE the United States |
| **Research articles published in peer-reviewed journals** | Review articles, position papers, case reports, commentary, editorials, personal narratives, conference abstracts, and unpublished works |

The full text for the remaining papers were then sourced and examined in detail with the complete list of inclusion and exclusion criteria (Table 2). Two reviewers (R1, R2) filtered remaining irrelevant papers from the review. Conflicts were discussed between both reviewers and resolved, and the remaining papers were included for later data extraction and synthesis. A flow chart detailing the screening process was created following the PRISMA guidelines and is shown in Figure 1.^10^

**Figure 1.**
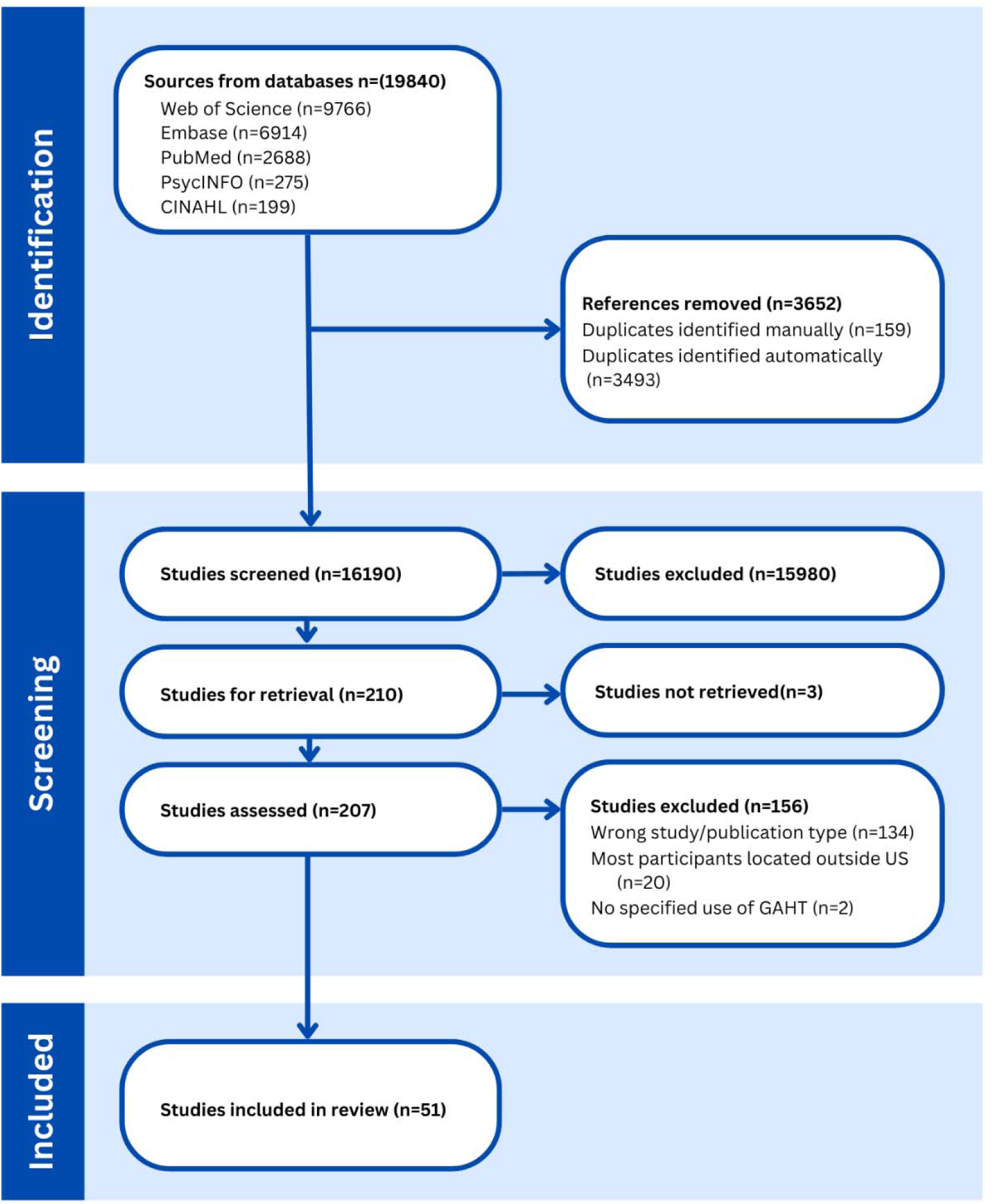
PRISMA Screening Flow-Chart from paper selection conducted for literature review on GAHT impact on quality of life.

### Data Extraction and Quality Assessment

Data extraction and quality assessment of the selected papers were completed using forms created by the study team in the Covidence software. The data extraction form was created referencing the University of North Carolina Health Sciences Library guide for systematic reviews and the Joanna Briggs Institute (JBI) Manual for Evidence Synthesis guidelines for scoping reviews.^11,12^ Pertinent information regarding participant demographics, study designs, study limitations, and main outcomes was recorded using this form.

A quality assessment tool based on the Mixed Methods Appraisal Tool was created in Covidence for quality assessment of the selected papers that included the evaluation of both quantitative and qualitative results.^13^ Two reviewers (R1,R2) independently completed the data extraction and quality assessment forms for all included papers, and conflicts were discussed and resolved. A composite quality assessment score was calculated for each paper by converting responses from a “no/maybe/yes” format to a numerical scale of “-1/0/1” for each assessment criteria. Papers received a score of “-1” if they did not meet the criteria, “0” if the criteria were deemed inapplicable or if reviewers were unable to determine if the criteria were met, and “1” if the criteria were fully met. Statistical comparison of quality assessment scores by publication date was assessed using a Pearson coefficient correlation test. The forms used for both processes are provided in Supplemental Materials B.

### Data Synthesis

To assess quality and quantity of information related to transgender healthcare as well as identify research gaps in the effects of gender-affirming care on well-being of TGNC patients, collected information was qualitatively assessed in a thematic analysis. This was achieved by assessing papers from five different aspects: participant characteristics, study designs and methods, quality assessment, recurring themes, and limits/barriers to studies. These aspects were qualitatively analyzed to assess for risk of bias in population representation, type of data, and quality of data.

## RESULTS

The search yielded 19,482 results across five databases, and 3,652 duplicates were identified and removed. The paper titles and abstracts were then screened for relevance and an additional 15,980 results were excluded. The full texts of the remaining 210 papers were assessed and further filtered to a final 51 papers that were included in data extraction and quality assessment. A PRISMA flow chart including the number of papers and specified reasons for exclusion is shown in Figure 1. A table of key extracted data is shown in Supplemental Table C.

### Descriptive Assessment

#### Participant Characteristics

The representation of different age groups varied across the reviewed papers with many papers including participants across several age groups. Young adults (18-30 years) were the most frequently represented age group, with 49 (96%) papers reporting data for this population. Middle-aged adults (31- 50 years) were represented in 31 (61%) papers, older adults (51+ years) were represented in 25 (49%) papers, and adolescents (10-17 years) were the least frequently reported, with 21 (41%) papers. These results are visualized in Figure 3.

Papers including TGNC people across a spectrum of gender identities, including nonbinary and gender-nonconforming individuals, accounted for 34 (67%) of the reviewed papers. Eight (16%) papers focused exclusively on binary transgender identities (female-to-male and male-to-female), while five (10%) papers were specific to female-to-male transgender individuals, and four (8%) papers focused solely on male-to-female transgender individuals.

Participant selection varied across studies highlighting the differences in stages of care under investigation. Studies required participants to be currently receiving or initiating care in eight (16%) papers, while 18 (35%) papers limited participants to those who were current patients at specific clinics. Three (6%) papers incorporated views from parents and caregivers of TGNC youth, two (4%) papers focused on the experiences of transgender veterans, and one (2%) study sought the perspectives of healthcare providers on issues related to transgender healthcare. Researchers provided financial compensation to participants in 27 (53%) papers.

#### Study Design and Methodology

Publication and data collection on this research topic has increased over time. Data collection was less prevalent in earlier decades, with only one (2%) study each during the 1980s and 1990s, and four (8%) studies conducted between 2000 and 2009. There has been an increase in research activity in recent decades. Between 2010 and 2019, data was collected for 42 (82%) papers, and 16 (31%) papers reported data collection occurring in 2020 or later. The publication dates for the included papers are represented graphically in Figure 4.

The geographical distribution of papers spanned all areas of the United States, but the number of papers representing each region was unequally distributed. Of the 51 reviewed papers, 24 (47%) papers adopted a broad scope, including TGNC people from across the United States. Regions represented were most prominent in the Northeast with eight (15%) papers and the West with 11 (22%) papers. The Southwest was represented by five (10%) papers and the Midwest by four (8%) papers, while the Southeast and non-contiguous states were each represented by a single study (2%). These results are mapped in Figure 2.

**Figure 2.**
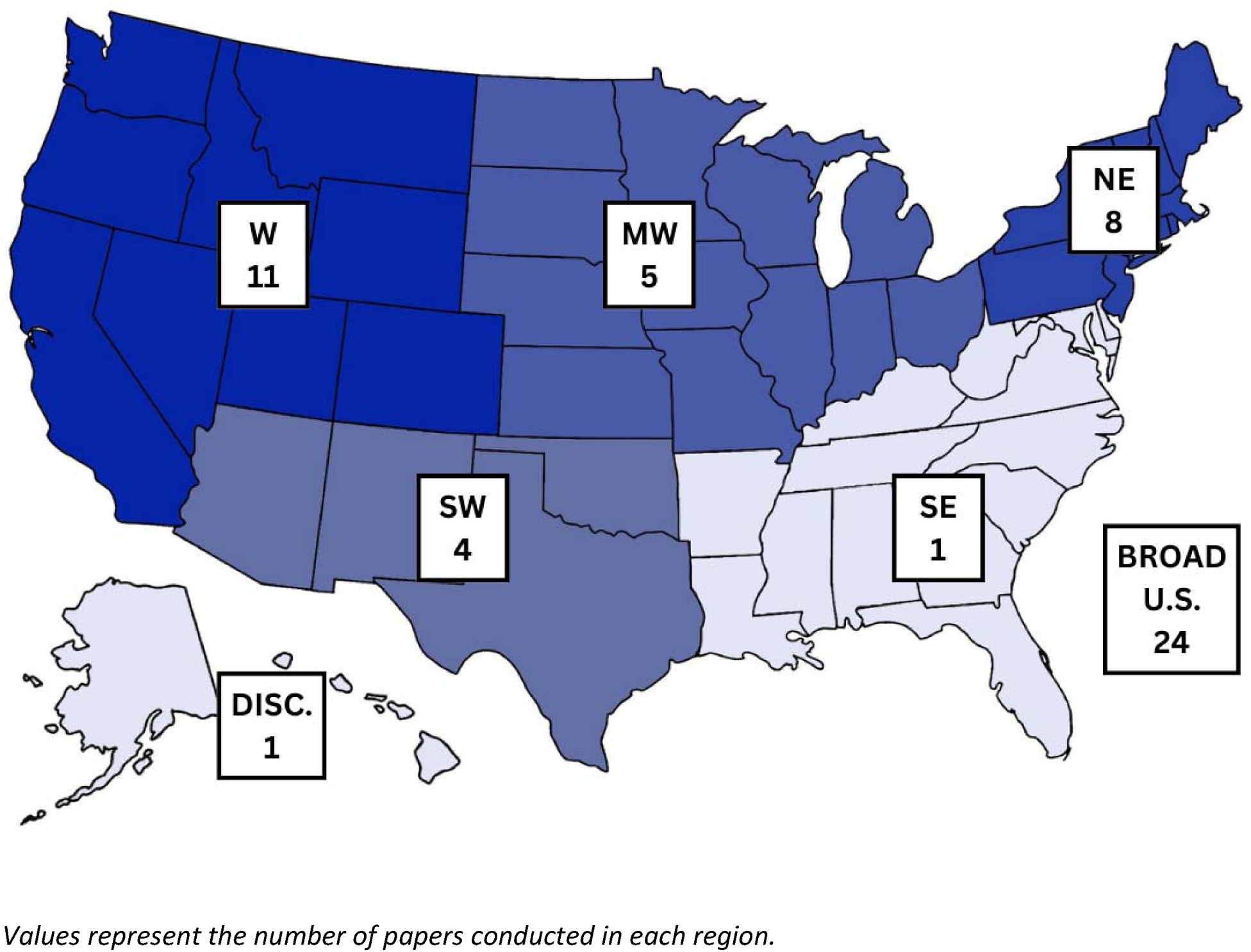
US map of number of papers by geographical region in study on GAHT impact on quality of life, (1984-2024)

**Figure 3.**
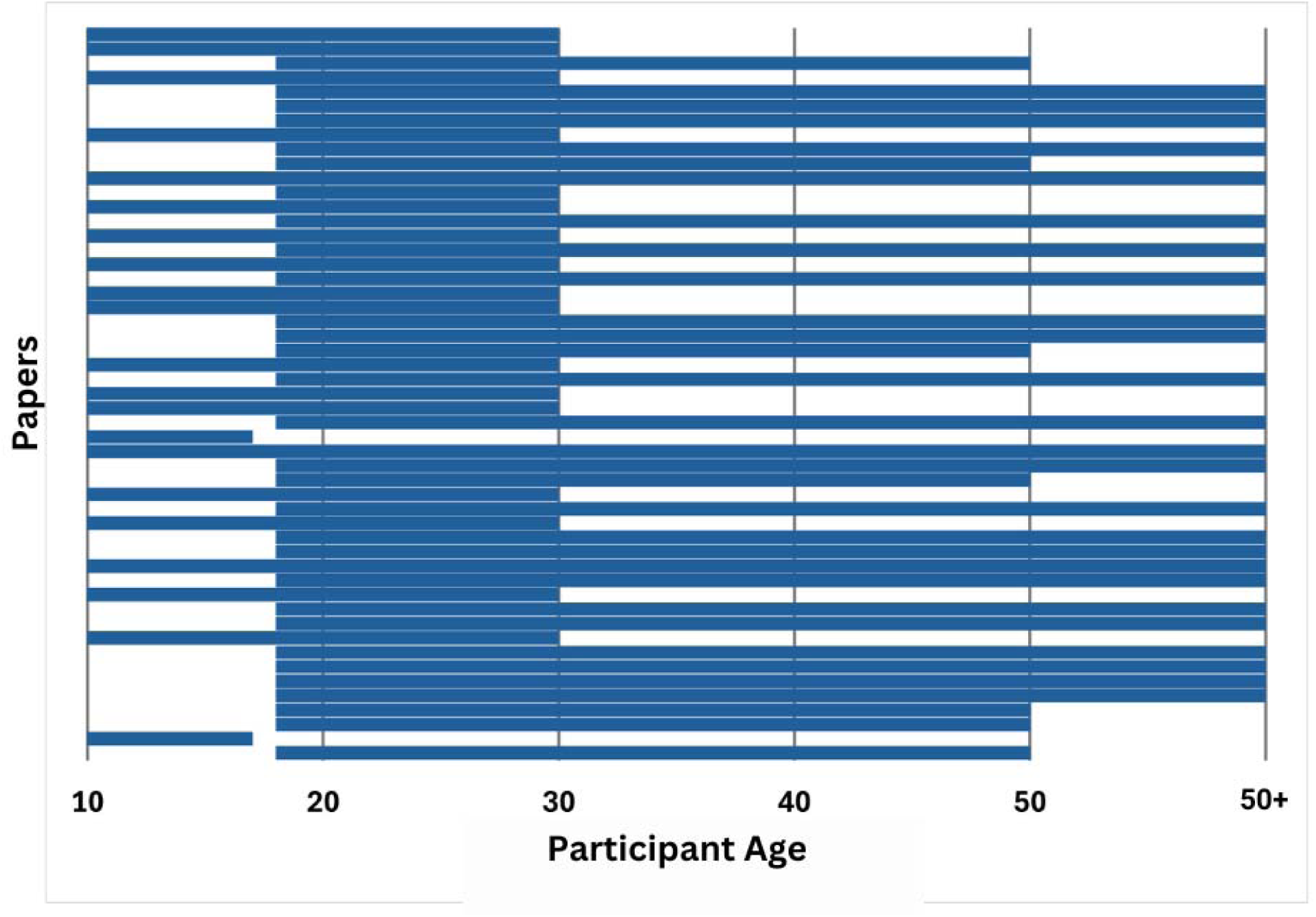
Age ranges of study participants in studies included in narrative review on GAHT impact on quality of life, (1984-2024)

**Figure 4.**
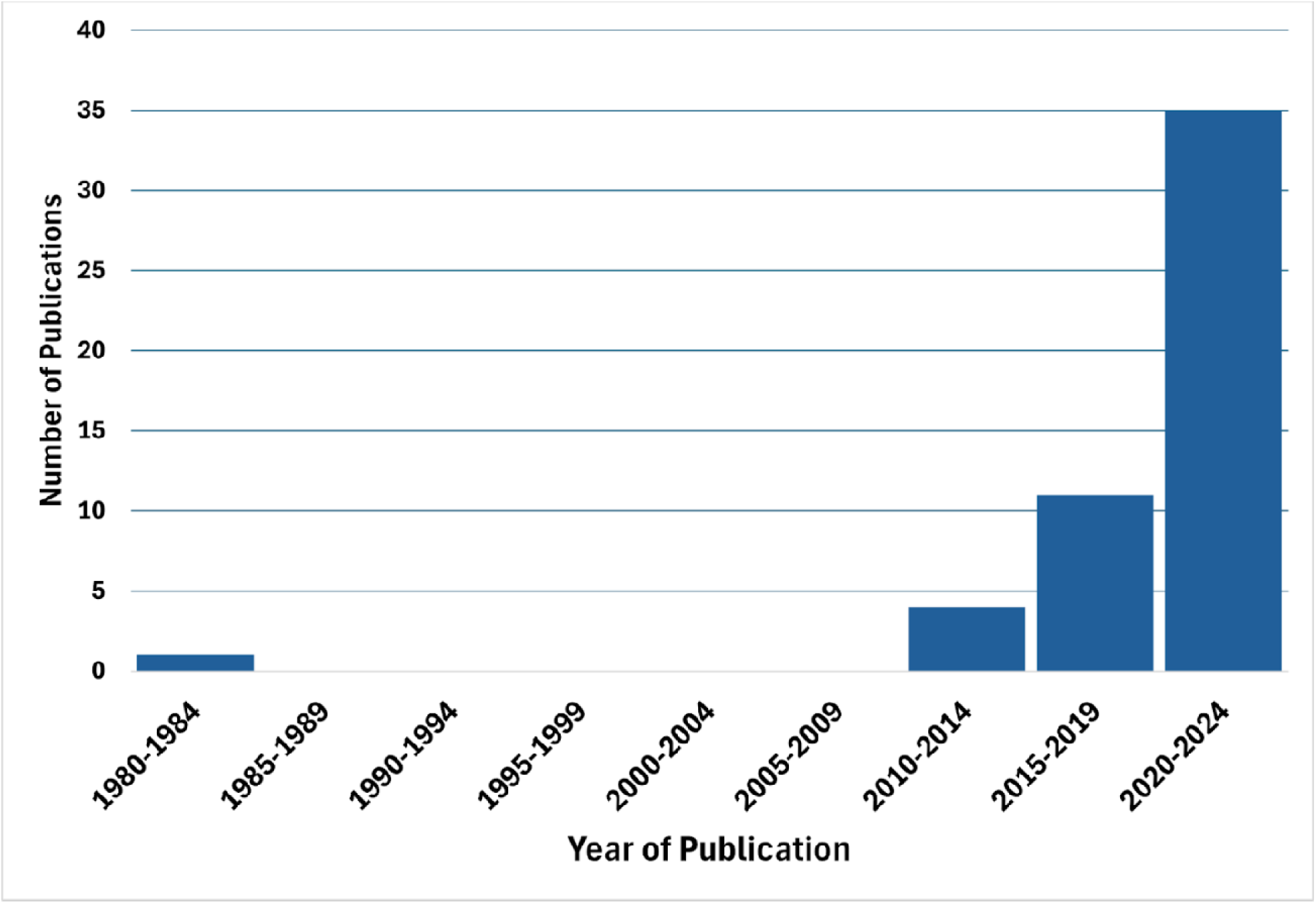
Publication dates of included papers in narrative review on GAHT impact on quality of life, (1984-2024)

A variety of study designs were used in the reviewed papers. As represented in Figure 5, 29 (57%) papers used cross-sectional designs, 13 (25%) papers used cohort designs, and seven (14%) adopted a qualitative approach. Randomized control trials and non-randomized experimental designs included one (2%) study each. Cohort designs were more frequently used with pediatric participants, with 10 (48%) papers involving adolescent participants utilizing this longitudinal approach.

**Figure 5.**
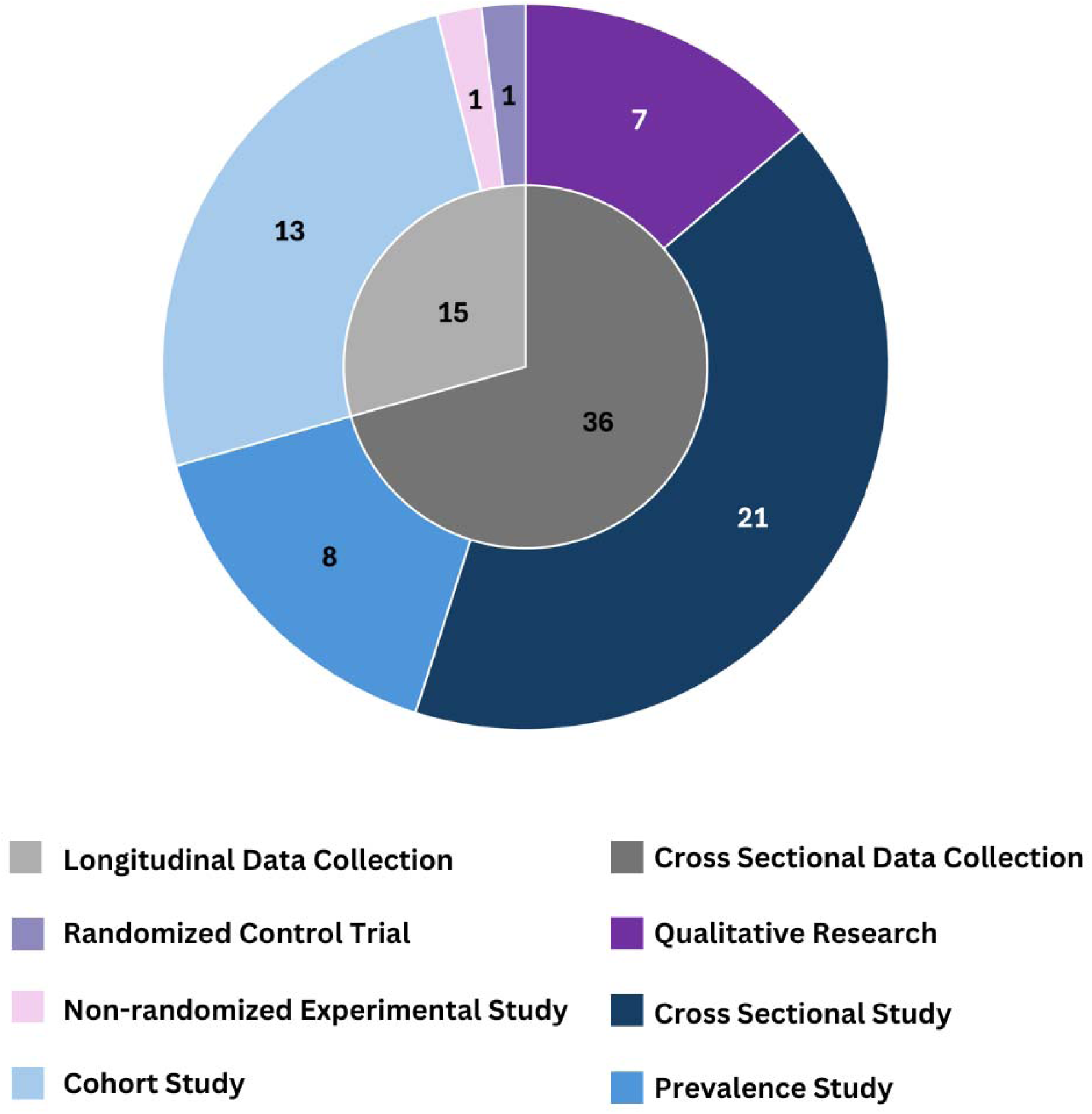
Study designs used in reviewed papers in narrative review on GAHT impact on quality of life, (1984-2024)

Papers reported different recruitment methods to identify and enroll study participants. The most frequently used sampling method involved recruiting participants from local clinics or LGBTQ+ affirming healthcare centers. Additional recruitment strategies included posting on online forums, social media platforms, and support groups. The majority of papers (26, 51%) reported recruiting with a single sampling method.

Methods of data collection varied among papers. Surveys were the primary method of data collection, used in 33 (65%) papers, followed by validated or published questionnaires, used in 27 (53%) papers. A total of 58 distinct scales were identified. The most frequently used tools were the Patient Health Questionnaire (PHQ-9)^14^ in six (11%) papers, the General Anxiety Disorder Scale (GAD-7)^15^ in five (10%) papers, and the Kessler-6 Psychological Distress Scale^16^ in five (10%) papers. In pediatric-inclusive papers, the most common tools were the PHQ-9 used in four (8%) papers and the GAD-7, Screen for Child Anxiety Related Disorders (SCARED)^17^, and Multidimensional Scale of Perceived Social Support (MSPSS)^18^ each used in three (5%) papers. Additionally, 10 of the identified scales were specifically designed for pediatric and adolescent populations. Interviews were conducted in six (12%) papers, and nine (18%) papers used other prospective data collection approaches. Publicly available data was sourced for 12 (24%) papers, while retrospective chart reviews were employed in 10 (20%) papers.

Most studies did not follow participants over long periods of time. The majority of included papers used a cross-sectional design, assessing participants at a single point in time with no longitudinal follow-up. In contrast, a smaller group of papers followed participants longitudinally, with three (6%) papers tracking participants for less than one year, eight (16%) papers following participants for one year, and two (4%) papers extending follow-up to two years. Only one (2%) study reported a follow-up period exceeding two years. The lengths of follow-up periods are represented graphically in Figure 6.

**Figure 6.**
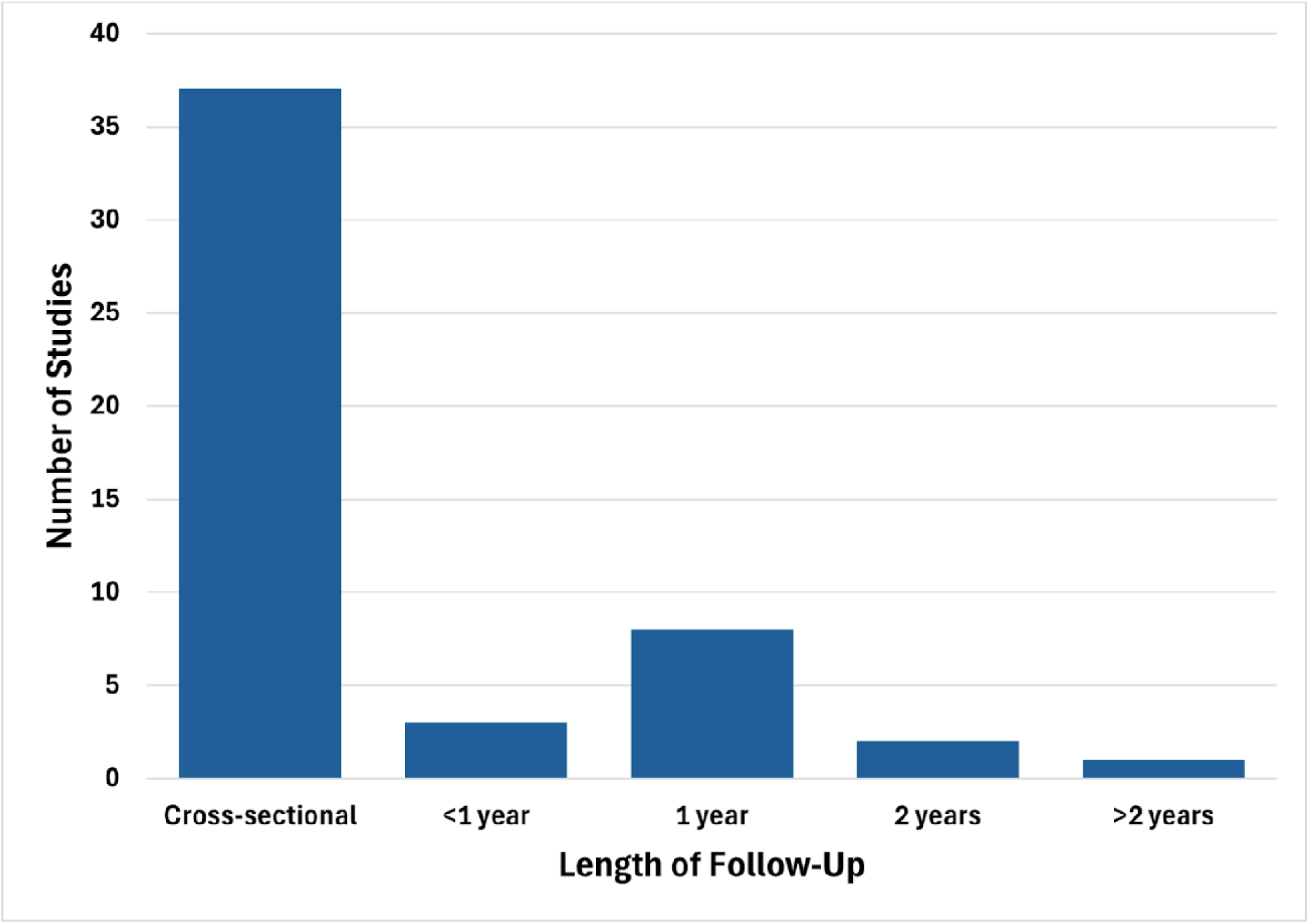
Lengths of follow-up reported in reviewed papers in narrative review on GAHT impact on quality of life, (1984-2024)

### Quality Assessment

The average quality assessment score on a scale ranging from −21 to 21 across all papers was 11.7. Scores ranged from −13, representing the poorest quality, to 19, the highest quality. The criteria least often met were: “Is there a significant risk of nonresponse bias?” with an average score of −0.51, “Were the study groups similar at the start of the study?” with an average score of 0, and “Would the intervention provide greater value compared to other interventions?” with an average score of 0.12. The Pearson correlation test comparing quality assessment scores to publication dates to assess whether the quality of research has improved over time yielded a non-significant coefficient of determination (R² = 0.38). Further inspection revealed the three papers with the lowest ratings were not associated with a specific period, being published in 1984, 2013, and 2020. The total scores for each study can be found in Supplemental Graph D.

### Thematic Analysis

The recurring themes across the included papers focused on two main topics: the mental health and quality of life outcomes for TGNC people receiving gender-affirming medical interventions and the disparities and barriers to accessing this care.

Multiple psychosocial outcomes were commonly measured across studies. Regularly measured outcomes included general mental health/wellbeing, suicidal thoughts or ideations, depression, anxiety, quality of life, distress, and alcohol/drug use. The most frequently assessed psychosocial outcomes were changes in depression, anxiety, self-harm, and overall well-being in relation to gender-affirming care.

Other outcomes of interest included levels of dysphoria, life satisfaction, appearance congruence, psychological distress, executive function, and substance use. Several papers reported significant psychosocial outcomes related to GAHT use. Overall, negative mental health indicators such as levels of depression, anxiety, and suicidal ideation were reported to be lower among individuals who had received GAHT. Suicidal ideation was reported to be lower for individuals receiving GAHT in 12 (24%) papers, while depression scores were lower in 11 (22%) papers, anxiety was lower in 10 (20%) papers. Positive mental health indicators such as quality of life and general mental health were reported to be higher among individuals who had received GAHT. Both quality of life and general mental health was higher in six (12%) papers. No statistically significant psychosocial changes were observed with treatment in five papers (10%).

The most commonly reported barriers to receiving gender-affirming care included the cost of treatment, lack of insurance coverage, limited access to competent providers, anticipated and experienced stigma or discrimination, insufficient education on standards of care for GAHT, geographic distance to clinics, state-level care bans, and general delays or limitations in access to care. A conceptual map depicting these themes is shown in Figure 7.

**Figure 7.**
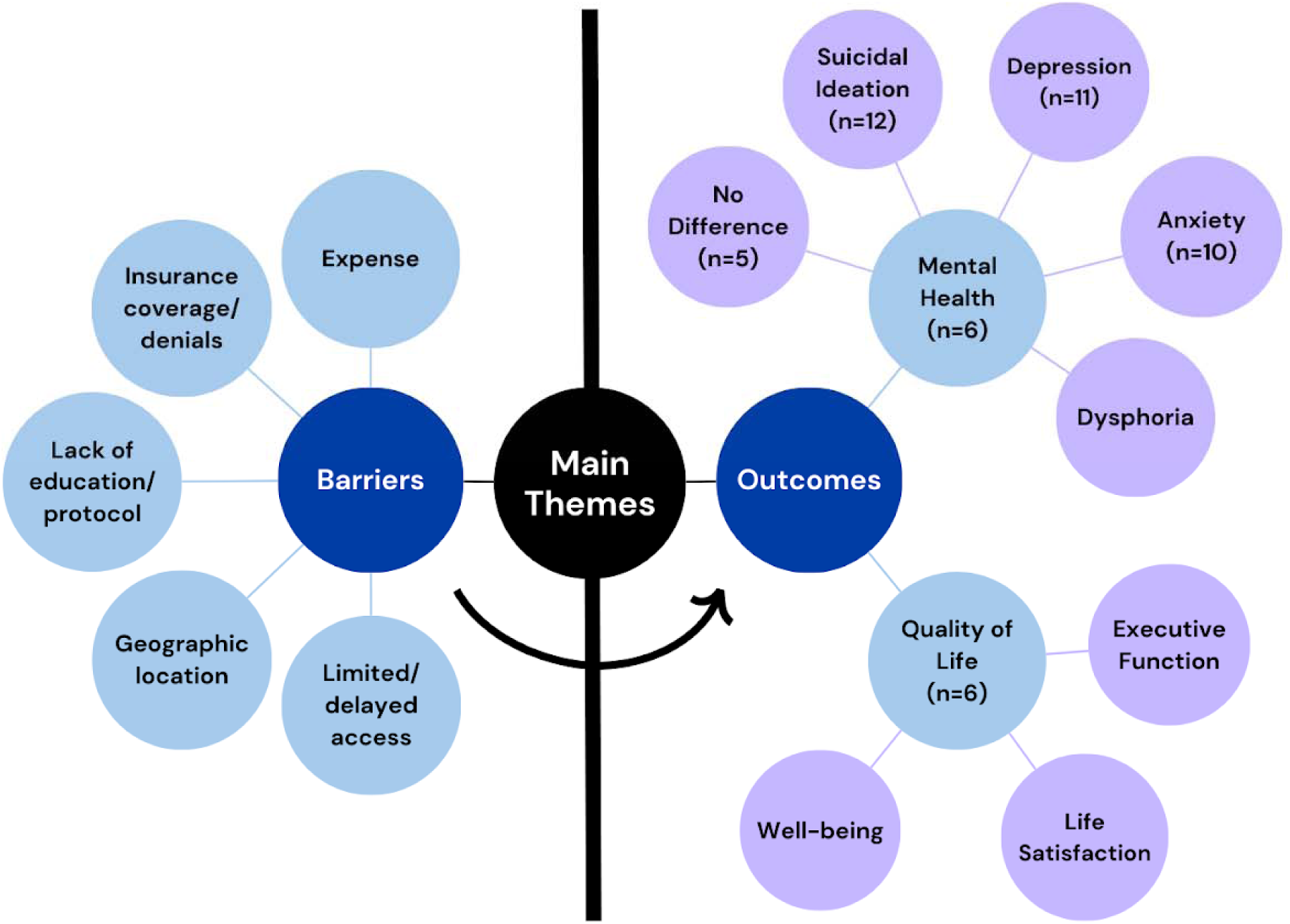
Conceptual map representation of main themes identified in narrative review on GAHT impact on quality of life, (1984-2024)

## DISCUSSION

The purpose of this narrative review was to evaluate the current evidence of the psychosocial impact of GAHT and identify gaps in knowledge to guide future research. We examined the relationship between TGNC patients seeking hormone replacement therapy and resulting improved health outcomes as well as the barriers they face in receiving this care.

### Descriptive Assessment

#### Participant Characteristics

Participant ages included in the papers identify a disproportionately higher representation of young adults, likely reflecting the many TGNC people that begin gender-affirming medical interventions in early adulthood. The relatively limited focus on younger adolescent and older adult age groups highlights the need for more inclusive age representation in future studies. Previous studies note that both groups would benefit from additional research as adolescents face unique developmental, social, and legal considerations in accessing care, while older adults encounter distinct challenges related to aging and age-related stigma.^19,20^

Our investigation also revealed a lack of research including nonbinary and other gender-nonconforming identities. Their inclusion in research is necessary, as nonbinary individuals often face unique healthcare disparities and needs that differ from those of binary transgender individuals.^21^ Additionally, identity-specific research, such as papers focused solely on male-to-female transgender individuals, are also important as they provide valuable insight into the specific needs and challenges associated with particular transition processes and experiences. All of these perspectives and scopes of research contribute to the development of individualized and adaptable care models that better support the diverse spectrum of TGNC identities.

The criteria for participant selection used in the reviewed papers often introduced a selection bias toward participants who have the resources and support to seek, obtain, and afford gender-affirming care. Demographics of participants tended to be more representative of white, educated, higher-income TGNC people residing in urban areas. This likely leads to the underrepresentation of several groups within the TGNC community such as people living in rural areas, people with lower socioeconomic statuses, and people from racial and ethnic minority groups that face additional barriers to accessing gender-affirming care and being included in research. Adolescents are also at risk of underrepresentation, as most require parental or caretaker consent to access GAHT. TGNC youth who do not have supportive family environments are thus less likely to be included in research requiring hormone use and have additionally been noted to have worse mental health outcomes than their peers with parental support.^22^ Furthermore, the high cost of puberty blockers in the United States necessitates financial stability or health insurance coverage, excluding economically disadvantaged youth from participation.^23^

Additional perspectives were included in five of the reviewed papers, adding another layer to the understanding of transgender healthcare. Parent and caregiver experiences, such as those provided by Gridley and colleagues,^24^ Kidd and colleagues,^25^ and Sequeria and colleagues,^26^ assist in understanding barriers to care for TGNC youth, transgender veterans by Tucker and colleagues^27^ and Wolfe and colleagues^28^ provide insight into distinct challenges such as navigating the Veterans Health Administration (VHA), and healthcare providers by Wolfe and colleagues can assist in identifying gaps in education and training as well as barriers that affect the quality of care provided to TGNC people. These diverse viewpoints help to establish more inclusive and effective healthcare practices and policies for all TGNC patients.

#### Study Design and Methodology

The increase in the number of papers conducted on GAHT in recent years reflects a growing recognition of the importance of research on gender-affirming care. After the first paper was published in 1984, no other papers were published that met the search criteria for this review until 2011. The limited number of papers from 1980-2010 highlights a historical gap in literature, which impeded earlier understanding of the health needs of TGNC people. The continuous rise in papers published from 2010 to today marks a shift in attention toward TGNC people and their health in both the social and scientific world. Papers that include pediatric participants have followed a similar trend, with the earliest study in this review being published in 2012.^29^ This increase in research activity provides a stronger evidence base to inform standards of care and ensure better health outcomes for the TGNC community.

The geographic distribution of the reviewed papers is more representative of people living in the Northeast and Western United States with fewer papers conducted in the Southeast and non-contiguous states. This pattern is partially reflected by the larger population of TGNC people in states such as California and New York, as well as the relatively progressive social and political climates in states like Oregon and Washington.^30,31^ Indeed, one paper specifically examined access and outcomes of GAHT in the state of Oregon because of the progressive legislation and insurance coverage.^32^ Florida additionally has a large population of TGNC people, but has a recent history of enacting anti-transgender legislation that limits access to gender-affirming healthcare, which may influence the availability and capacity of research conducted in the state.^30,33^ However, TGNC people reside and seek care across all regions of the United States, regardless of population or local policy. The underrepresentation of papers from the Southeast and non-contiguous states highlights a need for more research in these areas to improve health inequities for TGNC people.

Our search with this narrative review identified research conducted across the full range of exploratory to confirmatory methods, including qualitative studies and one randomized controlled trial on GAHT administration methods.^34^ The predominant number of cross-sectional papers are well-suited to identify associations between gender-affirming care and social and mental health outcomes, yet they are limited in their ability to establish causality or track changes with time. Longitudinal studies, although less common among the papers included in this review, can provide a more comprehensive understanding of the effects of gender-affirming care over time and are more useful in establishing causality. Papers with longer follow-up periods are particularly useful in providing critical data on the sustained effects of GAHT, supplying evidence related to the persistence of psychological, social, and physical outcomes. Qualitative research is often useful in identifying the complexities of emotional and social factors that influence and are influenced by gender-affirming care. These factors are often difficult to quantify, making qualitative approaches essential for a deeper understanding of the diversity of TGNC individuals’ lived experiences. The interpretation and application of qualitative findings also require caution as participant samples used in these studies are often inherently small and context-specific, making generalizability of results to broader populations limited. Finally, ethical concerns surrounding withholding treatment along with feasibility constraints in blinding due to the physical changes directly observable from hormone usage make it challenging to conduct randomized control trials, a gold standard in research.

Just over half of the included papers provided financial compensation for participant involvement. Offering compensation for a minority group’s participation in research is controversial. Financial compensation can act as an incentive to participants to enhance representation and participation within the study, which can contribute to ensuring that research findings are more generalizable and unbiased. Additionally, it acknowledges the time, effort, and potential risks undertaken by participants, fostering trust and reducing barriers to engagement between research and marginalized communities. On the other hand, there is concern for potential coercion into research participation with financial compensation, especially for those living in lower socioeconomic statuses.^35^ Participants should be able to freely consent to participation in research, with compensation fairly reflecting their time and effort while avoiding disproportionate targeting of lower-income communities.

The diversity of data collection methods in transgender health research reflects an intentional effort to represent both quantitative and qualitative aspects of TGNC experiences. Surveys were the most frequently noted data collection method and are efficient and versatile, but are prone to self-reporting bias, which affects the accuracy and reliability of participant responses. Chart reviews provide objective clinical data, offering the ability to track the use of healthcare and treatment outcomes, yet they are limited by the consistency and level of detail in available records, which potentially overlook social or other circumstantial factors. Interviews were used less frequently as they can be time-consuming, even for a small participant sample, but they offer insight into lived experiences that are often unobtainable through quantitative approaches alone. Finally, three different publicly available datasets were sourced for 12 papers, providing a time and cost-effective option for obtaining data. Repeated reliance on these data sets, however, lead to redundancy in findings and limits the ability to address newly emerging research questions. Continuing to employ diverse data collection methods will continue to reduce bias, increase generalizability, and offer a more complete view of transgender health.

Over half of the reviewed papers used validated or published scales to assess mental and social health outcomes. The large number of distinct scales identified highlights the complexity of mental health assessment and the absence of a universal standard for measuring psychological and social states. The use of scales specifically designed for pediatric and adolescent populations emphasize the importance of age-appropriate instruments that account for developmental and other contextual factors. While the variation in measurement tools allows for a comprehensive exploration of mental health, it complicates cross-study comparisons and meta-analyses. This highlights the need to standardize assessment practices to enhance comparability and ensure accurate evaluation of diverse populations.

### Quality Assessment

As this review included both qualitative and quantitative papers, we created a simplified quality assessment tool based on the Mixed Methods Appraisal Tool to assess scientific rigor and study quality across a broad range of research. The quality ratings in general were moderate to high, with only two (4%) papers scoring negative totals (Supplemental Graph D). While quality ratings were not significantly associated with a specific period, we found that the three highest-rated papers were published in 2023 and 2024, indicating a possible trend toward more rigorous standards in future transgender health research. This trend may be a response to increased demands for higher-quality research in this field. Nonresponse bias was a concern in most papers, as participants were often limited to individuals with the resources, access, and ability to seek and afford gender-affirming care. This limitation risks underrepresenting key groups within the TGNC community, such as those living in rural areas, individuals with lower socioeconomic status, and racial or ethnic minority groups, who face additional barriers to accessing care. The next lowest-scoring question, “Were the study groups similar at the start of the study?”, was often not applicable since many papers lacked comparison groups. Lastly, papers rarely evaluated interventions against alternatives such as withholding treatment, therapy, or surgical options. Two studies did compare outcomes from different GAHT administration methods.^34,36^ To improve future research, efforts should focus on reducing nonresponse bias by employing more inclusive recruitment strategies that prioritize underrepresented groups within the TGNC community. Studies should also aim to include diverse and comparable study groups where feasible, enabling more comparative analyses. Finally, researchers should aim to design studies that explicitly evaluate the relative effectiveness and value of various treatments or interventions, which would provide insight for optimizing care in this population.

### Thematic Analysis

Frequently identified themes in the included papers related to mental health and quality of life outcomes as well as barriers to accessing care aligns with much of the previously published research on transgender healthcare.

Findings surrounding psychosocial outcomes identify GAHT as having an essential role in enhancing mental health and social outcomes for TGNC people. Positive mental health indicators such as quality of life and general mental health were reported to be higher among individuals that received GAHT, while negative mental health indicators such as levels of depression, anxiety, and suicidal ideation were reported to be lower. No statistically significant changes were observed in a small selection of papers. Of note, only one paper of the 51 included reported a single statistically significant negative effect on psychosocial functioning. Strang and colleagues identified a relationship between long-term pubertal suppression and poorer executive function outcomes, but were unable to identify whether this was due to direct effects of puberty blockers or if longer periods of their use indicated youth were not deemed “ready to progress” to gender-affirming hormone replacement therapy for a variety of reasons.^37^ These mixed findings highlight the benefits of GAHT and related interventions for many individuals but also emphasizes the need for additional research to understand the factors that contribute to variability in individual outcomes.

The large number of financial, logistical, and legal barriers affecting TGNC patients reported in the papers is particularly concerning as these can restrict access to necessary medical care and exacerbate healthcare inequities, disproportionately affecting marginalized groups within the TGNC community. Barriers like high costs and limited provider availability may prevent early intervention, potentially leading to worse health outcomes later in life.^38^ Additionally, experiences of stigma or discrimination in healthcare settings can undermine trust in medical institutions, discouraging individuals from seeking care once it becomes accessible.^39^ Identifying and understanding these obstacles enables the development of strategies and policies to reduce disparities and promote equal access to healthcare. Changes such as increasing provider training, expanding insurance coverage for gender-affirming treatments, and advocating for legal protections against discriminatory practices are essential steps toward improving access.

### Limitations and Barriers

The reported limitations and barriers identified in the reviewed papers should be considered when interpreting findings, as they affect the strength of the conclusions and generalizability of the results to larger populations and diverse contexts. Commonly cited limitations within the papers included selection biases related to participant demographics, limited follow-up duration, and the lack of control groups for comparisons. Studies conducted online additionally contribute to these issues, as they inherently exclude individuals without reliable internet access. Individuals who were unable to access gender-affirming care due to financial, logistical, or legal barriers were rarely included, leaving substantial gaps in understanding the experiences of these underserved populations. Additional factors such as the COVID-19 pandemic and increasing anti-transgender legislation likely influenced findings in complex ways. These external variables have affected mental health and access to gender-affirming care, altering the observed relationships between gender-affirming care and psychosocial health outcomes.^40,41^

Limitations within this narrative review are also present. The initial collection of 19,482 papers was substantially reduced to 51 papers for data extraction and synthesis. This intentionally broad search strategy approach was deemed essential to ensure the strength of our findings and to mitigate potential limitations. The diverse, and in some cases outdated, terminology used to identify and affirm TGNC people across decades of publication required this methodological approach, allowing for the capture of a broad range of papers addressing the topics of this review. The rapidly growing body of research on transgender healthcare resulted in the exclusion of newer papers, as sourcing for this project ended in May 2024. Additionally, the data extraction and quality assessment relied on two researchers, which introduces bias based on individual perspectives and lived experiences. This project also focused specifically on the relationship between GAHT and psychosocial health. Papers that combined GAHT with other forms of gender-affirming care, such as therapy or surgery, were included in the review, but limit the ability to draw conclusions about the effects of GAHT alone.

### Future Research

To further advance the understanding of transgender health and improve quality of care for TGNC patients, this review identified gaps that future research should prioritize: longitudinal study designs with longer follow-up periods, control groups, and diverse participant bases. Additional longitudinal and prospective studies are needed to follow the long-term impacts of gender-affirming care on mental health, quality of life, and other related health outcomes. These studies allow for a better understanding of how frequently changing societal views and policies influence the health and well-being of TGNC people over time. Randomized controlled trials and mixed-methods designs should be used to build on the findings of the qualitative and exploratory studies reviewed here to strengthen causal inferences and explore more complex aspects of transgender health. Increasing sample sizes and including control groups for comparison, when possible, will additionally improve the quality of results.

Current research on transgender healthcare provides valuable knowledge surrounding mental health and gender-affirming care experiences of TGNC people, and there are many opportunities to improve this knowledge through more diverse, rigorous, and longitudinal approaches. This continued work is necessary for shaping future standards of care and policies that promote improved healthcare and outcomes for TGNC people. While research has seen greater focus on transgender healthcare and outcomes, many studies continue to lack diversity in participant populations, which limits the external validity of findings within the broader TGNC community. Future projects should prioritize including diverse participant groups including individuals from rural areas, lower socioeconomic statuses, and those with limited family or social support. The findings from the reviewed papers identified many barriers to accessing care. Geographic, economic, and systemic factors are frequently identified as obstacles to timely and affordable access to the current standards of care for TGNC patients. Addressing these barriers will improve clinical practice and reduce health disparities.

## CONCLUSION

The results of this narrative review emphasize the need for additional research on health outcomes and disparities in the TGNC community that focus on diverse participant populations and longitudinal study designs. This review additionally reinforces existing evidence supporting the importance of access to gender affirming care for TGNC individuals’ health and quality of life. With additional research and policy changes encouraging the development of improved holistic care for TGNC patients, we expect improved quality of care, patient satisfaction, and health outcomes for the TGNC community.

## Supporting information

Supplemental Graph D

## Data Availability

All data produced in the present study are available upon reasonable request to the authors

## ACKNOWLEDGEMENTS

We would like to thank Peter Johnson and A.R. Dykes Library of the Health Sciences at the University of Kansas Medical Center for their support in accessing the included databases and developing a search strategy.

Research reported in this publication was supported by the National Center for Advancing Translational Sciences of the National Institutes of Health under the Award Number UL1TR002366 to Rebecca Lepping. The content is solely the responsibility of the authors and does not necessarily represent the official views of the National Institutes of Health or the University of Kansas Medical Center.

## Notes

### Competing Interest Statement

The authors have declared no competing interest.

