## Supplemental Material A for "Gender-affirming hormone therapy and impacts on quality of life: a narrative review"

A. **Full search strategies used in literature review**

**PubMed**

("gender affirming hormon*" [tiab] OR “gender affirming hormon* therapy” [tiab] OR “hormone replacement therapy” [tiab] OR “HRT” [tiab] OR “gender affirming treatment” [tiab] OR "GAHT" [tiab] OR "gender diverse hormone" [tiab] OR "hormon* treatment*" [tiab] OR "hormon* therapy" [tiab] OR “gender-affirming care” [tiab] OR “puberty inhibitors” [Mesh] OR “puberty inhibitors” [tiab] OR “puberty blockers” [tiab] OR "Hormone Antagonists" [Mesh] OR “transgender healthcare” [tiab] OR “androgens” [Mesh] OR “estrogens” [Mesh] OR “estrogen*” [tiab] OR “testosterone” [Mesh] OR “testosterone” [tiab] OR “cross sex hormon*” [tiab] OR “CSH” [tiab] OR "Gonadal Steroid Hormones" [Mesh] OR “feminizing hormon*” [tiab] OR “FHT” [tiab] OR “feminizing treatment” [tiab] OR “masculinizing hormon*” [tiab] OR “MHT” [tiab] OR “masculinizing treatment” [tiab] OR "Steroid Synthesis Inhibitors" [Mesh] OR “testosterone block*” [tiab] OR “Endocrinology” [Mesh] OR “endocrinology” [tiab] OR “Endocrine treatment” [tiab] OR "Health Services for Transgender Persons" [Mesh])

AND

(LGBT* [tiab] OR GLBT* [tiab] OR LGBQ* [tiab] OR LGBS* [tiab] OR M2F [tiab] OR GLBQ* [tiab] OR GLBs* [tiab] OR 2SLGBT* [tiab] OR “two-spirit” [tiab] OR GBMSM* [tiab] OR msm [tiab] OR TGNC* [tiab] OR YTW [tiab] OR transgender* [tiab] OR transpeople* [tiab] OR "men loving men" [tiab] OR "men who have sex with men" [tiab] OR "women loving women" [tiab] OR "women who have sex with women" [tiab] OR transvestite [tiab] OR "cross sex" [tiab] OR crosssex* [tiab] OR crossgender* [tiab] OR F2M [tiab] OR FTM [tiab] OR MTF [tiab] OR MtX [tiab] OR FtX [tiab] OR “pangender” [tiab] OR “third gender” [tiab] OR transperson* [tiab] OR hijra* [tiab] OR transsexual* [tiab] OR intersex* [tiab] OR queer* [tiab] OR genderqueer [tiab] OR agender [tiab] OR demigender [tiab] OR "gender queer" [tiab] OR "trans* people" [tiab] OR "trans* individual" [tiab] OR "trans* individuals" [tiab] OR "trans* person" [tiab] OR "trans* persons" [tiab] OR "trans* sexual" [tiab] OR "trans* man" [tiab] OR "trans* men" [tiab] OR "trans* male" [tiab] OR “transmen” [tiab] OR “transmasculine” [tiab] OR “trans masculine” [tiab] OR “transfeminine” [tiab] OR “trans feminine” [tiab] OR “transwomen” [tiab] OR "trans* women" [tiab] OR "trans* woman" [tiab] OR "trans* female" [tiab] OR "trans* youth" [tiab] OR “trans* adolescent” [tiab] OR "trans* population" [tiab] OR "trans* gender" [tiab] OR "sexual minority" [tiab] OR "sexual minorities" [tiab] OR dysphoria [tiab] OR “gender dysphori*” [tiab] OR “gender euphori*” [tiab] OR "transitioned people" [tiab] OR "transitioned person" [tiab] OR "transitioned population" [tiab] OR “gender transition” [tiab] OR “gender non-conforming” [tiab] OR “gender-nonconforming” [tiab] OR "Sexual and Gender Minorities" [Mesh] OR "Transsexualism" [Mesh] OR "Gender-Nonconforming Persons" [mh] OR "Intersex Persons" [mh] OR "Transgender Persons" [mh] OR “transgender person*” [tiab] OR "Gender Identity" [Mesh] OR “gender identity” [tiab] OR “Gender Diverse People” [tiab] OR “gender divers*” [tiab] OR “nonbinary” [tiab] OR “enby” [tiab] OR “non-binary” [tiab] OR “gender reassignment” [tiab] OR “gender incongruen*” [tiab] OR “gender identity disorder” [tiab] OR “female to male” [tiab] OR “male to female” [tiab] OR “transgender identity” [tiab] OR “genderfluid” [tiab] OR “genderqueer” [tiab])

AND

("patient compliance" [mh] OR "patience compliance" [tiab] OR "patient adherence" [tiab] OR "Patient Satisfaction" [Mesh] OR "patient satisfaction" [tiab] OR "Quality of Life" [Mesh] OR "quality of life" [tiab] OR "life quality" [tiab] OR "Psychological Well-Being" [mh] OR "well being" [tiab] OR "patient experience" [tiab] OR "cultural competency" [mh] OR "cultural competenc*" [tiab] OR "Treatment Outcome" [Mesh] OR “treatment outcome” [tiab] OR "Mental Health" [Mesh] OR “mental health” [tiab] OR “behavioral health” [tiab] OR “behavioral wellness” [tiab] OR “mood” [tiab] OR “psychological effects” [tiab] OR “self-esteem” [tiab] OR “mental distress” [tiab] OR "Delivery of Health Care" [Mesh] OR "Long-Term Care" [Mesh] OR "Health Inequities" [Mesh] OR "Quality of Health Care" [Mesh] OR "Health Services Accessibility" [Mesh])

**PsycINFO and CINAHL**

(TI "gender affirming hormon*" OR AB "gender affirming hormon*" OR TI “gender affirming hormon* therapy” OR AB “gender affirming hormon* therapy” OR TI “hormone replacement therapy” OR AB “hormone replacement therapy” OR TI “HRT” OR AB “HRT” OR TI “gender affirming treatment” OR AB “gender affirming treatment” OR TI "GAHT" OR AB "GAHT" OR TI "gender diverse hormone" OR AB "gender diverse hormone" OR TI "hormon* treatment*" OR AB "hormon* treatment*" OR TI "hormon* therapy" OR AB "hormon* therapy" OR TI “gender-affirming care” OR AB “gender-affirming care” OR TI “transgender healthcare” OR AB “transgender healthcare” OR TI “cross sex hormon*” OR AB “cross sex hormon*” OR TI “CSH” OR AB “CSH” OR TI “feminizing hormon*” OR AB “feminizing hormon*” OR TI “FHT” OR AB “FHT” OR TI “feminizing treatment” OR AB “feminizing treatment” OR TI “masculinizing hormon*” OR AB “masculinizing hormon*” OR TI “MHT” OR AB “MHT” OR TI “masculinizing treatment” OR AB “masculinizing treatment” OR TI “testosterone block*” OR AB “testosterone block*” OR TI “Endocrine treatment” [tiab] OR AB “Endocrine treatment” [tiab] OR TI “puberty blockers” OR AB “puberty blockers” OR DE “Hormone Therapy” OR DE “Gender Affirming Care” OR DE “Sex Hormones” OR DE “androgens” OR DE “testosterone” OR DE “antiandrogens” OR DE “antiestrogens” OR DE “gonadotropic hormones” OR DE “endocrinology”)

AND

(TI LGBT* OR AB LGBT* OR TI GLBT* OR AB GLBT* OR TI LGBQ* OR AB LGBQ* OR TI LGBS* OR AB LGBS* OR TI M2F OR AB M2F OR TI GLBQ* OR AB GLBQ* OR TI GLBs* OR AB GLBs* OR TI 2SLGBT* OR AB 2SLGBT* OR TI “two-spirit” OR AB “two-spirit” OR TI GBMSM* OR AB GBMSM* OR TI msm OR AB msm OR TI TGNC* OR AB TGNC* OR TI YTW OR AB YTW OR TI transgender* OR AB transgender* OR TI transpeople* OR AB transpeople* OR TI "men loving men" OR AB "men loving men" OR TI "men who have sex with men" OR AB "men who have sex with men" OR TI "women loving women” OR AB "women loving women” OR TI "women who have sex with women" OR AB "women who have sex with women" OR TI transvestite OR AB transvestite OR TI "cross sex" OR AB "cross sex" OR TI crosssex* OR AB crosssex* OR TI crossgender* OR AB crossgender* OR TI F2M OR AB F2M OR TI FTM OR AB FTM OR TI MTF OR AB MTF OR TI MtX OR AB MtX OR TI FtX OR AB FtX OR TI “pangender” OR AB “pangender” OR TI “third gender” OR AB “third gender” OR TI transperson* OR AB transperson* OR TI hijra* OR AB hijra* OR TI transsexual* OR AB transsexual* OR TI intersex* OR AB intersex* OR TI queer* OR AB queer* OR TI genderqueer OR AB genderqueer OR TI agender OR AB agender OR TI demigender OR AB demigender OR TI "gender queer" OR AB "gender queer" OR TI "trans* people" OR AB "trans* people" OR TI "trans* individual" OR AB "trans* individual" OR TI "trans* individuals" OR AB "trans* individuals" OR TI "trans* person" OR AB "trans* person" OR TI "trans* persons" OR AB "trans* persons" OR TI "trans* sexual" OR AB "trans* sexual" OR TI "trans* man" OR AB "trans* man" OR TI "trans* men" OR AB "trans* men" OR TI "trans* male" OR AB "trans* male" OR TI “transmen” OR AB “transmen” OR TI “transmasculine” OR AB “transmasculine” OR TI “trans masculine” OR AB “trans masculine” OR TI “transfeminine” OR AB “transfeminine” OR TI “trans feminine” OR AB “trans feminine” OR TI “transwomen” OR AB “transwomen” OR TI "trans* women" OR AB "trans* women" OR TI "trans* woman" OR AB "trans* woman" OR TI "trans* female" OR AB "trans* female" OR TI "trans* youth" OR AB "trans* youth" OR TI “trans* adolescent” OR AB “trans* adolescent” OR TI "trans* population" OR AB "trans* population" OR TI "trans* gender" OR AB "trans* gender" OR TI "sexual minority" OR AB "sexual minorit*" OR TI dysphoria OR AB dysphoria OR TI “gender dysphori*” OR AB “gender dysphori*” OR TI “gender euphori*” OR AB “gender euphori*” OR TI "transitioned people" OR AB "transitioned people" OR TI "transitioned person" OR AB "transitioned person" OR TI "transitioned population" OR AB "transitioned population" OR TI “gender transition” OR AB “gender transition” OR TI “gender non-conforming” OR AB “gender non-conforming” OR TI “gender-nonconforming” OR AB “gender-nonconforming” OR TI “Gender Diverse People” OR AB “Gender Diverse People” OR TI “gender divers*” OR AB “gender divers*” OR TI “nonbinary” OR AB “nonbinary” OR TI “enby” OR AB “enby” OR TI “non-binary” OR AB “non-binary” OR TI “gender reassignment” OR AB “gender reassignment” OR TI “gender incongruen*” OR AB “gender incongruen*” OR TI “gender identity disorder” OR AB “gender identity disorder” OR TI “female to male” OR AB “female to male” OR TI “transgender identity” OR AB “transgender identity” OR TI “gender identity” OR AB “gender identity” OR TI “gender expression” OR AB “gender expression” OR TI “genderfluid” OR AB “genderfluid” OR TI “genderqueer” OR AB “genderqueer” OR TI “male to female” OR AB “male to female” OR DE “transgender” OR DE “LGBTQ” OR DE “Gender expression” OR DE “gender reassignment” OR DE “gender transition” OR DE “sexual minority groups” OR DE “transsexualism” OR DE “gender nonconforming” OR DE “gender dysphoria” OR DE “gender identity” OR DE “intersex”)

AND

(TI "patience compliance" OR AB "patience compliance" OR TI "patient adherence" OR AB "patient adherence" OR TI "patient satisfaction" OR AB "patient satisfaction" OR TI "quality of life" OR AB "quality of life" OR TI "life quality" OR AB "life quality" OR TI "well being" OR AB "well being" OR TI "patient experience" OR AB "patient experience" OR TI "cultural competenc*" OR AB "cultural competenc*" OR TI “behavioral health” OR AB “behavioral health” OR TI “psychological effects” OR AB “psychological effects” OR TI “behavioral wellness” OR AB “behavioral wellness” OR TI “mood” OR AB “mood” OR TI “self-esteem” OR AB “self-esteem” OR TI “mental distress” OR AB “mental distress” OR TI “psychological well-being” OR AB “psychological well-being” OR TI “treatment outcome” OR AB “treatment outcome” OR TI “mental health” OR AB “mental health OR TI “delivery of health care” OR AB “delivery of health care” OR TI “health inequit*” OR AB “health inequit*” OR TI “quality of health care” OR AB “quality of health care” OR DE “treatment compliance” OR DE “medical regimen compliance” OR DE “patient adherence” OR DE “client attitudes” OR DE “treatment barriers” OR DE “treatment outcomes” OR DE “medical regimen compliance” OR DE “client satisfaction” OR DE “quality of life” OR DE “well being” OR DE “subjective well being” OR DE “cultural competence” OR DE “youth mental health” OR DE “mental health” OR DE “health care delivery” OR DE “quality of care” OR DE “quality of life”)

**Embase**

(‘gender affirming hormon*’:ti,ab OR ‘gender affirming hormon* therapy’:ti,ab OR ‘hormone replacement therapy’:ti,ab OR ‘HRT’:ti,ab OR ‘gender affirming treatment’:ti,ab OR ‘GAHT’:ti,ab OR ‘gender diverse hormone’:ti,ab OR ‘hormon* treatment*’:ti,ab OR ‘hormon* therapy’:ti,ab OR ‘gender-affirming care’:ti,ab OR ‘puberty inhibitors’:ti,ab OR ‘puberty blockers’:ti,ab OR ‘transgender healthcare’:ti,ab OR ‘estrogen*’:ti,ab OR ‘testosterone’:ti,ab OR ‘cross sex hormon*’:ti,ab OR ‘CSH’:ti,ab OR ‘feminizing hormon*’:ti,ab OR ‘FHT’:ti,ab OR ‘feminizing treatment’:ti,ab OR ‘masculinizing hormon*’:ti,ab OR ‘MHT’:ti,ab OR ‘masculinizing treatment’:ti,ab OR ‘testosterone block*’:ti,ab OR ‘endocrinology’:ti,ab OR ‘Endocrine treatment’:ti,ab OR 'hormonal therapy’/exp OR ‘gender reassignment’/exp OR ‘gender-affirming care’/exp OR ‘hormone antagonist’/exp OR ‘sex hormone’/exp OR ‘endocrinology’/exp)

AND

(LGBT*:ti,ab OR GLBT*:ti,ab OR LGBQ*:ti,ab OR LGBS*:ti,ab OR M2F:ti,ab OR GLBQ*:ti,ab OR GLBs*:ti,ab OR 2SLGBT*:ti,ab OR two-spirit:ti,ab OR GBMSM*:ti,ab OR msm:ti,ab OR TGNC*:ti,ab OR YTW:ti,ab OR transgender*:ti,ab OR transpeople*:ti,ab OR ‘men loving men’:ti,ab OR ‘men who have sex with men’:ti,ab OR ‘women loving women’:ti,ab OR ‘women who have sex with women’:ti,ab OR transvestite:ti,ab OR ‘cross sex’:ti,ab OR crosssex*:ti,ab OR crossgender*:ti,ab OR F2M:ti,ab OR FTM:ti,ab OR MTF:ti,ab OR MtX:ti,ab OR FtX:ti,ab OR ‘pangender’:ti,ab OR ‘third gender’:ti,ab OR transperson*:ti,ab OR hijra*:ti,ab OR transsexual*:ti,ab OR intersex*:ti,ab OR queer*:ti,ab OR genderqueer:ti,ab OR agender:ti,ab OR demigender:ti,ab OR ‘gender queer’:ti,ab OR ‘trans* people’:ti,ab OR ‘trans* individual’:ti,ab OR ‘trans* individuals’:ti,ab OR ‘trans* person’:ti,ab OR ‘trans* persons’:ti,ab OR ‘trans* sexual’:ti,ab OR ‘trans* man’:ti,ab OR ‘trans* men’:ti,ab OR ‘trans* male’:ti,ab OR ‘transmen’:ti,ab OR ‘transmasculine’:ti,ab OR ‘trans masculine’:ti,ab OR ‘transfeminine’:ti,ab OR ‘trans feminine’:ti,ab OR ‘transwomen’:ti,ab OR ‘trans* women’:ti,ab OR ‘trans* woman’:ti,ab OR ‘trans* female’:ti,ab OR ‘trans* youth’:ti,ab OR ‘trans* adolescent’:ti,ab OR ‘trans* population’:ti,ab OR ‘trans* gender’:ti,ab OR ‘sexual minority’:ti,ab OR ‘sexual minorities’:ti,ab OR dysphoria:ti,ab OR ‘gender dysphori*’:ti,ab OR ‘gender euphori*’:ti,ab OR ‘transitioned people’:ti,ab OR ‘transitioned person’:ti,ab OR ‘transitioned population’:ti,ab OR ‘gender transition’:ti,ab OR ‘gender non-conforming’:ti,ab OR ‘gender-nonconforming’:ti,ab OR ‘transgender person*’:ti,ab OR ‘gender identity’:ti,ab OR ‘Gender Diverse People’:ti,ab OR ‘gender divers*’:ti,ab OR ‘nonbinary’:ti,ab OR ‘enby’:ti,ab OR ‘non-binary’:ti,ab OR ‘gender reassignment’:ti,ab OR ‘gender incongruen*’:ti,ab OR ‘gender identity disorder’:ti,ab OR ‘female to male’:ti,ab OR ‘male to female’:ti,ab OR ‘transgender identity’:ti,ab OR ‘genderfluid’:ti,ab OR ‘genderqueer’:ti,ab OR ‘sexual and gender minority’/exp OR ‘gender identity’/exp OR ‘gender dysphoria’/exp OR ‘gender transition’/exp)

AND

(‘patient compliance’:ti,ab OR ‘patient adherence’:ti,ab OR ‘patient satisfaction’:ti,ab OR ‘quality of life’:ti,ab OR ‘life quality’:ti,ab OR ‘well being’:ti,ab OR ‘patient experience’:ti,ab OR ‘cultural competenc*’:ti,ab OR ‘treatment outcome’:ti,ab OR ‘mental health’:ti,ab OR ‘behavioral health’:ti,ab OR ‘behavioral wellness’:ti,ab OR ‘mood’:ti,ab OR ‘psychological effects’:ti,ab OR ‘self-esteem’:ti,ab OR ‘mental distress’:ti,ab OR ‘patient attitude’/exp OR ‘patient satisfaction’/exp OR ‘quality of life’/exp OR ‘mental health’/exp OR ‘cultural competence’/exp OR ‘treatment outcome’/exp OR ‘health care delivery’/exp OR ‘health disparity’/exp OR ‘health care quality’/exp OR ‘health care access’/exp)

**Web of Science**

((gender affirming hormon*) OR (gender affirming hormon* therapy) OR (hormone replacement therapy) OR (HRT) OR (gender affirming treatment) OR (GAHT) OR (gender diverse hormone) OR (hormon* treatment*) OR (hormon* therapy) OR (gender-affirming care) OR (puberty inhibitors) OR (puberty blockers) OR (Hormone Antagonists) OR (transgender healthcare) OR (androgens) OR (estrogen*) OR (testosterone) OR (cross sex hormon*) OR (CSH) OR (Gonadal Steroid Hormones) OR (feminizing hormon*) OR (FHT) OR (feminizing treatment) OR (masculinizing hormon*) OR (MHT) OR (masculinizing treatment) OR (Steroid Synthesis Inhibitors) OR (testosterone block*) OR (endocrinology) OR (Endocrine treatment) OR (Health Services for Transgender Persons))

AND

((LGBT*) OR (2SLGBT*) OR (two-spirit) OR (TGNC*) OR (YTW) OR (transgender*) OR (transpeople*) OR (transvestite) OR (FTM) OR (MTF) OR (MtX) OR (FtX) OR (transperson*) OR (transsexual*) OR (intersex*) OR (queer*) OR (genderqueer) OR (gender queer) OR (trans* people) OR (trans* individual*) OR (trans* person*) OR (trans* sexual) OR (trans* man) OR (trans* men) OR (trans* male) OR (transmen) OR (transmasculine) OR (trans masculine) OR (transfeminine) OR (trans feminine) OR (transwomen) OR (trans* women) OR (trans* woman) OR (trans* female) OR (trans* youth) OR (trans* adolescent) OR (trans* population) OR (trans* gender) OR (sexual minorit*) OR (gender dysphori*) OR (transitioned people) OR (transitioned person) OR (transitioned population) OR (gender transition) OR (gender non-conforming) OR (gender-nonconforming) OR (Gender Minorities) OR (transgender person*) OR (gender identity) OR (gender divers*) OR (nonbinary) OR (non-binary) OR (gender reassignment) OR (gender incongruen*) OR (gender identity disorder) OR (female to male) OR (male to female) OR (transgender identity) OR (genderfluid) OR (genderqueer))

AND

((patience compliance) OR (patient adherence) OR (patient satisfaction) OR (quality of life) OR (life quality) OR (well being) OR (patient experience) OR (cultural competenc*) OR (treatment outcome) OR (mental health) OR (behavioral health) OR (behavioral wellness) OR (mood) OR (psychological effects) OR (self-esteem) OR (mental distress) OR (Delivery of Health Care) OR (Long-Term Care) OR (Health Inequities) OR (Quality of Health Care) OR (Health Services Accessibility))
