## Supplemental Material B for "Gender-affirming hormone therapy and impacts on quality of life: a narrative review"

B. **Data extraction and quality assessment forms used in literature review**

**GAHT Data Extraction**

Title	Authors

_________________________________	_________________________________

Study ID	Publication location (journal, website, etc)

_________________________________	_________________________________

Year of publication	Notes (any other study identification info)

_________________________________	_________________________________

**Study Characteristics**

**Demographics of participants**

Country of Residence	Reported Age Range

   o Only United States	  ◻ Adolescent: 10-17

   o US + Canada	 	◻ Young adult: 18-30

   o US + Other(s)	  ◻ Middle-aged: 31-50

   o Not reported	  ◻ Older adults: 50+

   o Other: __________________

Gender Identity

Region(s) of US	 	o FTM only

West: WA, OR, CA, NV, ID, MT, WY, UT, CO	  o MTF only

Southwest: AZ, NM, OK, TX	  o FTM + MTF only

Midwest: ND, SD, NE, KS, MN, IA, MO, WI,	  o All TGD and GNC identities

IL, MI, IN, OH	  o Other: __________________

Southeast: AR, LA, MS, AL, GA, FL, TN, KY,

SC, NC, VA, WY, MD, DE	Race

Northeast: ME, NH, MA, CT, NJ, PA, NY, RI, VT	 	◻ American Indian or Alaska Native

   ◻ West	  ◻ Asian

   ◻ Midwest	  ◻ Black or African American

   ◻ Southwest	  ◻ Native Hawaiian or Pacific Islander

   ◻ Southeast	◻ Hispanic or Latino

   ◻ Northeast	  ◻ White

   ◻ Discontiguous states and territories	 	◻ Not reported

   ◻ Not reported	  ◻ Other: ___________________

Household Income/Socioeconomic Status	Ethnicity

   ◻ Lower: <$30,000	  ◻ Hispanic or Latino

   ◻ Lower-middle: $30,000-58,000	  ◻ Not Hispanic or Latino

   ◻ Middle: $58,001-94,000	  ◻ Not reported

   ◻ Upper-middle: $94,001-153,000

   ◻ Upper: >$153,000	Insurance Coverage

   ◻ Not reported	  ◻ Private insurance

  ◻ Public insurance

Sexual Orientation	  ◻ No insurance

   ◻ Straight	  ◻ Not reported

   ◻ Lesbian

   ◻ Gay	Total Number of Participants

   ◻ Bisexual	___________________________

   ◻ Pansexual

   ◻ Asexual	Any other demographic information

   ◻ Not reported	___________________________

   ◻ Other: __________________

Inclusion Criteria	Exclusion Criteria

___________________________	___________________________

___________________________	___________________________

___________________________	___________________________

___________________________	___________________________

**Study Details**

Study Design	Participant sampling methods

   o Randomized control trial	  ◻ Snowball sampling

   o Non-randomized experimental study	  ◻ Social media advertising

   o Cohort study	  ◻ Flyer advertisement

   o Cross sectional study	  ◻ Clinic recruitment

   o Case control study	 	◻  Support groups

   o Qualitative research	  ◻  Online forums

   o Prevalence study	  ◻  Other: ___________________

   o Other: __________________

Therapies reported

Participants Compensated	  ◻ Pre-therapy (if desired)

   o Yes	 	◻ Puberty suppressors/blockers

   o No	  ◻ Hormone replacement therapy

   ◻ HRT + surgery

Control group (if applicable)	  ◻ Puberty blockers + HRT

___________________________	  ◻ Puberty blockers + HRT + surgery

   ◻ Surgery

   ◻ Other:_____________________

Follow up interval(s) (if applicable)	Study Setting

___________________________	  o Independent clinic

  o Hospital

Period of Data Collection	  o Community setting

   ◻ 1980-1989	 	o Online

   ◻ 1990-1999	  o Other

   ◻ 2000-2009

   ◻ 2010-2019	Source of Public Data (if applicable)

   ◻ 2020+	  ◻ 2015 US Transgender Survey

  ◻ Trans Health Survey

Method of Data Collection	  ◻ Trans Stress and Health Study

   ◻ Validated/published questionnaire	 	◻ Behavioral Risk Factor Surveillance

   ◻ Survey	  ◻ Other: ___________________

   ◻ Interview

   ◻ Public data	Any other study details

   ◻ Chart review	________________________________

   ◻ Other: __________________

Published Scales/Inventories/Questionnaires

   ◻ Ask Suicide Screening Questions (ASQ)	◻ Beck Anxiety Inventory

   ◻ Beck Depression Inventory - II	◻ CDC Youth Risk Behavior

   ◻ Behavior Rating Inventory of Executive	◻ Child and Adolescent Symptom Inventory

       Function (BRIEF)	    (CASI-5)

   ◻ Depression, Anxiety, and Stress Scale	◻ Flanagan Quality of Life Scale (QOLS)

       (DASS)	◻ GAD-7

   ◻ General Well-Being Scale (GWBS)	◻ Kessler-6 Psychological Distress Scale

   ◻ Mini-Social Phobia Inventory (Mini-SPIN)	◻ Patient-Reported Outcomes

   ◻ PHQ-2	    Measurement Information System

   ◻ PHQ-9	    (PROMIS)

   ◻ Quick Inventory of Depressive Symptoms	◻ Revised Children’s Manifest Anxiety

       (QIDS)	    Scale
   ◻ Rosenberg Self Esteem Scale	◻ Satisfaction With Life Scale

   ◻ Screen for Child Anxiety Related Emotional	◻ Short Form 36-Item Questionnaire

       Disorders (SCARED)	◻ Suicidal Behaviors Questionnaire (SBQ)

   ◻ Toolbox Emotion Battery	◻ WHO QOL

   ◻ Other(s): _________________________________________________________________

**Outcomes**

Results type

   o Quantitative

   o Qualitative

   o Mixed

Outcomes Measured (Please list all outcomes)

   ◻ General mental illness	◻ General mental health/wellbeing

   ◻ Depression	◻ Anxiety/social anxiety

   ◻ Quality of life	◻ Satisfaction with life

   ◻ Positive affect	◻ Self-esteem

   ◻ Executive functioning	◻ Suicidal thoughts/ideation

   ◻ Attempted suicide	◻  Self-harm

   ◻ Other(s): ____________________________________________________________

Please list 5 most relevant outcomes

**Outcome 1**

Outcome measured	Statistical Significance

______________________________	______________________________

Type	Confidence Interval

   o Quantitative	______________________________

   o Qualitative

   o Mixed

Outcome

   o Improved

   o No change

   o Worsened

   o Unable to tell

   o Other:______________________

Other outcomes measured

_______________________________________________________________________

**Limits to Study**

Barriers/Limits to study	Self-Reported Quality of Data

_______________________________	_______________________________

_______________________________	_______________________________

_______________________________	_______________________________

Any other outcome or barrier info

_______________________________

_______________________________

**Quality Assessment**

Did the study address a clearly focused research question?

Yes	No	Can’t tell

Was the methodology appropriate for the research question?

Yes	No	Can’t tell

Has the relationship between researcher and participants been adequately considered?

(Could the researcher’s identity or relationship with the participant group impact the way data is collected and interpreted?)

Yes	No	Can’t tell

Was the recruitment strategy appropriate to the aims of the research?

Yes	No	Can’t tell

Are specific inclusion/exclusion criteria used?

Yes	No	Can’t tell

Were all participants who entered the study accounted for at its conclusion?

Yes	No	Can’t tell	N/A

Were the study groups similar at the start of the study?

Yes	No	Can’t tell	N/A

Do the benefits of the intervention outweigh the harms and costs?

Yes	No	Can’t tell

Was the follow up of participants complete enough?

(Was enough data collected at each interval to contribute to answering the research question?)

Yes	No	Can’t tell	N/A

Was the follow up of participants long enough?

(Were participants followed long enough to report a noticeable effect of the therapy?)

Yes	No	Can’t tell	N/A

Were all variables and interventions clearly described?

Yes	No	Can’t tell

Can the results be applied/generalized to other populations/contexts easily?

(Can results be generally applied to other trans individuals in the US?)

Yes	No	Can’t tell

Would the intervention provide greater value compared to other interventions?

Yes	No	Can’t tell

Is there a clear statement of findings?

Yes	No	Can’t tell

Were the data analysis methods appropriate?

Yes	No	Can’t tell

Was the statistical significance/confidence interval given?

Yes	No	Can’t tell	N/A

Are confounding variables accounted for in the design and analysis?

Yes	No	Can’t tell

Is there a significant risk of nonresponse bias?

Yes	No	Can’t tell

Is the interpretation of results sufficiently substantiated by data?

Yes	No	Can’t tell

Do the results of this study fit with other available evidence?

Yes	No	Can’t tell

Have ethical issues been taken into consideration?

Yes	No	Can’t tell
