## Supplemental Table C for "Gender-affirming hormone therapy and impacts on quality of life: a narrative review"

C. **Data extraction summary from search conducted for review on GAHT impact on quality of life, (1984-2024)**

|  | **Study ID** | **Study Design** | **Sample size** | **Age Range** | **Gender Identities** | **Location** | **Other Demographics Reported** |
| --- | --- | --- | --- | --- | --- | --- | --- |
| 1 | Garcia 2020^32^ | Qualitative Research | 25 | 18-50 | MTF | West USA | Race, Sexual Orientation, Insurance Coverage |
| 2 | Sequeria 2023^26^ | Qualitative Research | 33 | 10-17 | All TGNC | West USA | Race, Ethnicity |
| 3 | Zatloff 2021^42^ | Qualitative Research | 27 | 18-50 | All TGNC | Southeast USA | Race |
| 4 | Turban 2020^43^ | Cross Sectional Study | 27,715 | 18-50 | All TGNC | USA + Territories | Race, Income, Sexual Orientation |
| 5 | Reisner 2023^44^ | Cross Sectional Study | 2,192 | 18-50+ | All TGNC | Northeast USA | Race, Ethnicity, Insurance Coverage |
| 6 | Tomita 2019^45^ | Cross Sectional Study | 1,427 | 18-50+ | FTM + MTF | USA | Race, Ethnicity, Income |
| 7 | Wilson 2015^46^ | Cross Sectional Study | 314 | 18-50+ | MTF | West USA | Race, Ethnicity, Income, Insurance Coverage |
| 8 | Hughes 2022^47^ | Cross Sectional Study | 11,994 | 18-50+ | All TGNC | USA | Race, Ethnicity, Insurance Coverage |
| 9 | Kidd 2023^25^ | Cross Sectional Study | 277 | 10-30 | All TGNC | USA | Race, Ethnicity |
| 10 | Kelly 2023^48^ | Cross Sectional Study | 101 | 18-50+ | All TGNC | USA | Race |
| 11 | Hughto 2020^49^ | Prevalence Study | 288 | 18-50+ | All TGNC | USA | Race |
| 12 | Reisner 2015^50^ | Cohort Study | 360 | 10-30 | FTM + MTF | Northeast USA | Race, Ethnicity |
| 13 | Lee 2024^51^ | Cross Sectional Study | 27,715 | 18-50+ | All TGNC | USA + Territories | Race, Ethnicity, Sexual Orientation, Insurance Coverage |
| 14 | Puckett 2022^52^ | Cross Sectional Study | 861 | 10-50+ | All TGNC | Not reported | Race, Ethnicity, Income, Sexual Orientation |
| 15 | Strenth 2023^53^ | Cross Sectional Study | 311 | 18-50+ | FTM + MTF | Southwest USA | Race, Insurance Coverage |
| 16 | Kattari 2020^54^ | Prevalence Study | 20,921 | 18-50+ | All TGNC | USA + Territories | Race, Ethnicity, Income |
| 17 | Gadomski 2023^55^ | Cohort Study | 173 | 10-30 | All TGNC | Northeast USA | Race, Ethnicity |
| 18 | Mann 2024^56^ | Prevalence Study | 6,598 | 18-50+ | All TGNC | USA + Territories | Race, Income, Sexual Orientation, Insurance Coverage |
| 19 | Allen 2019^57^ | Cohort Study | 47 | 10-30 | All TGNC | Midwest USA | Race, Ethnicity, Income, Insurance Coverage |
| 20 | Johnson 2022^58^ | Cross Sectional Study | 99 | 18-50 | All TGNC | USA | Race, Ethnicity, Income, Sexual Orientation |
| 21 | ColtonMeier 2011^59^ | Prevalence Study | 369 | 18-50+ | FTM | USA + Other Country | Race, Ethnicity, Income |
| 22 | Meier 2014^29^ | Non-randomized Experimental Study | 233 | 10-50+ | FTM | Southwest USA + Canada | Race, Ethnicity |
| 23 | Hobson 2022^23^ | Qualitative Research | 4 | 10-17 | All TGNC | Northeast USA | Race, Ethnicity, Income, Sexual Orientation |
| 24 | Klein 2023^60^ | Cross Sectional Study | 27,715 | 18-50+ | All TGNC | USA + Territories | Race, Ethnicity |
| 25 | Tordoff 2023^61^ | Cohort Study | 113 | 10-30 | All TGNC | West USA | Race, Ethnicity, Income, Insurance Coverage |
| 26 | Cantu 2020^62^ | Cohort Study | 80 | 10-30 | All TGNC | West USA | Not Reported |
| 27 | Goetz 2023^63^ | Qualitative Research | 54 | 18-50+ | All TGNC | USA + Canada | Race, Ethnicity |
| 28 | Achille 2020^64^ | Cohort Study | 116 | 10-30 | FTM + MTF | Northeast USA | Not Reported |
| 29 | Cooper 1984^65^ | Cohort Study | 16 | 18-50 | MTF | Discontiguous states or territories | Race, Ethnicity |
| 30 | Turban 2022^66^ | Cross Sectional Study | 27,715 | 18-50+ | All TGNC | USA + Territories | Race, Ethnicity, Income, Sexual Orientation |
| 31 | Tucker 2018^27^ | Cross Sectional Study | 312 | 18-50+ | FTM + MTF | USA | Race, Ethnicity, Income, Sexual Orientation |
| 32 | Strang 2022^37^ | Cross Sectional Study | 131 | 10-30 | All TGNC | Midwest and Northeast USA | Race, Ethnicity |
| 33 | Kuper 2020^67^ | Cohort Study | 209 | 10-30 | All TGNC | Southwest USA | Race, Ethnicity, Sexual Orientation |
| 34 | Stroumsa 2020^68^ | Prevalence Study | 26,957 | 18-50+ | All TGNC | USA + Territories | Race, Ethnicity, Insurance Coverage |
| 35 | Chelliah 2024^22^ | Cohort Study | 156 | 10-30 | All TGNC | Southwest USA | Race, Ethnicity, Sexual Orientiation |
| 36 | deHaan 2015^69^ | Cross Sectional Study | 314 | 18-50+ | MTF | West USA | Race, Ethnicity, Income |
| 37 | Moyer 2019^70^ | Cross Sectional Study | 194 | 10-30 | All TGNC | West USA | Not Reported |
| 38 | Wolfe 2023^28^ | Qualitative Research | 55 | 18-50+ | All TGNC | USA | Race, Ethnicity, Sexual Orientation |
| 39 | Green 2022^71^ | Cross Sectional Study | 11,914 | 10-30 | All TGNC | USA | Race, Ethnicity, Sexual Orientation |
| 40 | Bahr 2024^36^ | Cohort Study | 112 | 18-30 | MTF | Midwest USA | Race, Ethnicity |
| 41 | Gridley 2016^24^ | Qualitative Research | 65 | 10-50+ | All TGNC | West USA | Race, Ethnicity |
| 42 | Staples 2020^72^ | Cross Sectional Study | 396 | 18-50 | All TGNC | USA | Race, Ethnicity, Income |
| 43 | Butler 2019^73^ | Prevalence Study | 715 | 18-50+ | FTM + MTF | USA + Canada | Race, Ethnicity, Income |
| 44 | Baines 2023^34^ | Randomized control trial | 26 | 10-30 | FTM | West USA | Race, Ethnicity |
| 45 | Bakko 2020^74^ | Prevalence Study | 11,320 | 18-50+ | All TGNC | USA | Race, Ethnicity, Income, Insurance Coverage |
| 46 | Leinung 2013^75^ | Cohort Study | 242 | 18-50+ | FTM + MTF | Northeast USA | Not Reported |
| 47 | Cai 2019^20^ | Prevalence Study | 2,420 | 18-50+ | FTM + MTF | USA | Race, Ethnicity, Income, Insurance Coverage |
| 48 | Olsavsky 2023^76^ | Cross Sectional Study | 82 | 10-30 | All TGNC | Midwest USA | Race, Ethnicity |
| 49 | Watt 2018^77^ | Cross Sectional Study | 134 | 18-50 | FTM | Not reported | Race, Ethnicity |
| 50 | Chen 2023^78^ | Cohort Study | 315 | 10-30 | All TGNC | West, Midwest, and Northeast USA | Race, Ethnicity |
| 51 | Tordoff 2022^79^ | Cohort Study | 169 | 10-30 | All TGNC | West USA | Race, Ethnicity |
