## Supplementary figures and images for "Gender-affirming hormone therapy and impacts on quality of life: a narrative review"

### Supplemental Graph D

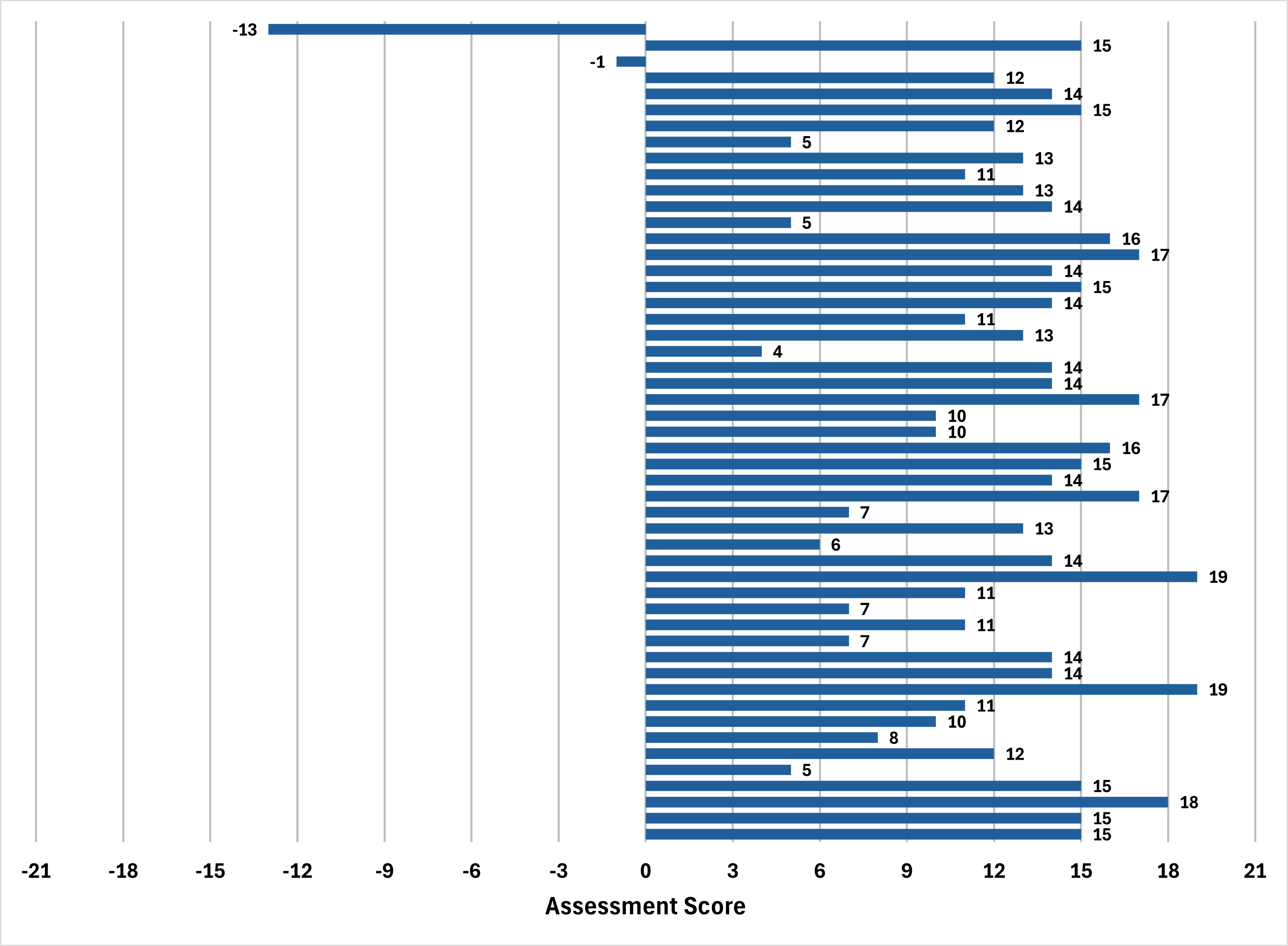
